# From clinics to sewers: leveraging environmental surveillance and whole genome sequencing to inform transmission of ESBL-*Escherichia coli* in Switzerland

**DOI:** 10.1101/2025.09.12.25335617

**Authors:** Sheena Conforti, Louis du Plessis, Claudia Bagutti, Jens Becker, Silvio D. Brugger, Alexia Cusini, Adrian Egli, Valeria Gaia, Gilbert Greub, Cornelia Guler, Jana S. Huisman, Claudine Kocher, Roger D. Kouyos, Karoline Leuzinger, Carola Maffioli, Mara Neacsu, Oliver Nolte, Alban Ramette, Salome N. Seiffert, Sarah Tschudin-Sutter, Jon Paulin Zumthor, Tanja Stadler, Timothy R. Julian

## Abstract

Extended-spectrum β-lactamase (ESBL)-producing *Escherichia coli* is a major antimicrobial resistance concern spreading across human, animal, and environmental domains. To assess between-source transitions, we analysed 762 ESBL-*E. coli* genomes collected from wastewater (used as a community shedding indicator), clinical settings, cattle, and wildlife across Switzerland (2021–2023). ST131 was the most prevalent sequence type (ST), and 76% of isolates carried resistance to at least two antibiotic classes in addition to β-lactams. Phylogenetic analysis showed isolates were interspersed across sources, yet genetically similar strains were more common within compartments. Clonal isolates (0 SNPs) were rarely shared (n = 2) between wastewater and corresponding clinics. Ancestral state reconstruction revealed compartmentalization of isolates between wastewater and clinics across the whole phylogeny. However, this pattern disappears within human-associated ST131, ST69, ST1193, highlighting exchange between clinics and communities. These findings show that wastewater surveillance captures community circulation of ESBL-*E. coli,* which overlap with circulating clinically-relevant strains.

## MAIN

In 2019, infections caused by extended-spectrum β-lactamase (ESBL)-producing *Escherichia coli* accounted for approximately 100,000 of the nearly 5 million deaths associated with bacterial antimicrobial resistance (AMR)(1). Initially predominant in hospitals, over the past 40 years ESBL-*E. coli* has spread to communities, livestock and wildlife (2, 3). Its widespread detection highlights the importance of not only human-to-human transmission, but also interactions within and between humans, livestock, and wildlife (3). Bacteria and antibiotic resistance genes (ARGs) exchange across these compartments, driving the spread of resistance through multiple transmission pathways (4–6). Accordingly, the World Health Organization (WHO) advocates a One Health approach for tackling AMR, emphasizing the interconnectedness of human, animal, and environmental health (6–8).

The environment (soil, water, wastewater, and hospital equipment) often acts as an intermediary in transmission, with humans and animals shedding ESBL-*E. coli* into it and being exposed through contact. The environment can also act as a reservoir where ESBL-*E. coli* can grow under suitable conditions, such as in soil (9, 10). Human activities that disturb natural habitats intensify inter-compartmental transmission, increasing AMR bacterial carriage in wildlife, while agricultural practices such as manure application contribute to ARGs cross-sectoral transmission (11, 12). Direct transmission from livestock to farmers can further expose humans to animal-derived resistance factors (4). Unlike many infectious diseases, the spread of AMR is driven not only by infection and colonization but also by horizontal gene transfer and long-term, asymptomatic carriage, both of which facilitate the persistence and dissemination of resistant organisms (13, 14). While there is evidence of cross-compartmental transmission events (15), understanding their direction and frequency may help designing effective interventions to reduce the spread of AMR.

Wastewater-based surveillance (WBS) is emerging as a useful tool for AMR monitoring within a One Health framework, tracking the presence of resistant bacteria in wastewater and providing insights into prevalence of both pathogenic and non-pathogenic resistant bacteria in communities (16–18). WBS offers a comprehensive view of AMR dissemination by capturing resistant bacteria from diverse sources, including sewer-connected entities like households, hospitals, and industries, as well as non-point sources like agricultural or urban runoff (19). The diversity of resistant bacteria from various sources can be further resolved using whole genome sequencing (WGS), which provides detailed insights into their distribution across human, animal, and environmental compartments, enhancing AMR surveillance (20). In the United Kingdom, the adoption of WGS for real-time surveillance of resistant pathogens resulted in enhanced public health outcomes by enabling quicker outbreak resolution, cost reduction, and increased confidence in linking sources to cases (21). Furthermore, WGS of wastewater isolates may help map transmission across compartments, contribute to epidemiological modelling, and guide public health strategies (16, 22).

In this study, we investigated the diversity and transmission of 762 ESBL-producing *E. coli* isolates across animal, clinical, and environmental compartments, including wastewater, in Switzerland to elucidate potential transmission pathways within a One Health framework. Specifically, we aimed to: *i)* characterize the genetic diversity of ESBL-producing *E. coli* across multiple compartments, *ii)* assess whether ESBL-producing *E. coli* diversity shows clustering based on source, geographic regions, or both, and *iii)* determine transition events between sources using phylogenomic approaches. Given that wastewater captures bacteria shed almost exclusively by human populations, we use wastewater isolates as an indicator for community-level ESBL-*E. coli*. Our findings provide insights into the diversity and transmission dynamics of ESBL-*E. coli* in Switzerland, contributing to a comprehensive AMR surveillance strategy at the national scale.

## RESULTS

### Sampling and sequencing

A total of 834 *Escherichia coli* isolates producing extended-spectrum β-lactamases (ESBL) were recovered from four sources across Switzerland between November 2021 and October 2023: wastewater (n = 438), healthcare facilities (n = 328), cattle (n = 26), and wildlife (n = 42) (**Fig. 1, Fig. S1, Extended Data Table 1**). Clinical isolates were obtained through a convenience sampling strategy, whereby only confirmed ESBL-producing *E. coli* from inpatients and outpatients at healthcare facilities were included. Of the 834 isolates sequenced, 72 (9%) were excluded due to incorrect species assignment and/or contamination (n = 45), low assembly quality (n = 21), or low sequencing coverage (n = 6) (**Table S1**). The remaining 762 (91%) passed quality control and were included in downstream analyses (**Table S2, Table S3**). Exclusions were relatively consistent across sources, with 406 of 438 (93%) wastewater, 302 of 328 (92%) clinical, 25 of 26 (96%) cattle, and 29 of 42 (69%) wildlife isolates retained. Isolates were distributed across sources as follows: 406 (53%) from wastewater, 302 (40%) clinical isolates from in/out-patients at healthcare facilities, 29 (4%) from wildlife and 25 (3%) from cattle (**Table S1**). To monitor contamination and sequencing performance, 12 negative extraction controls (AE blanks) were included across both sequencing batches, with no evidence of contamination detected. Reference strains of *E. coli* (strain 1 and 2; n = 8 and 8 in batch 1, n = 1 and 1 in batch 2) and *Klebsiella pneumoniae* (strain 1 and 2; n = 8 and 5 in batch 1, n = 1 and 2 in batch 2) were successfully sequenced across both batches with expected results. *E. coli* strains matched previously sequenced genomes, and *K. pneumoniae* strains were clearly distinguishable, confirming the accuracy of both sequencing and downstream species assignment.

**Fig. 1:**
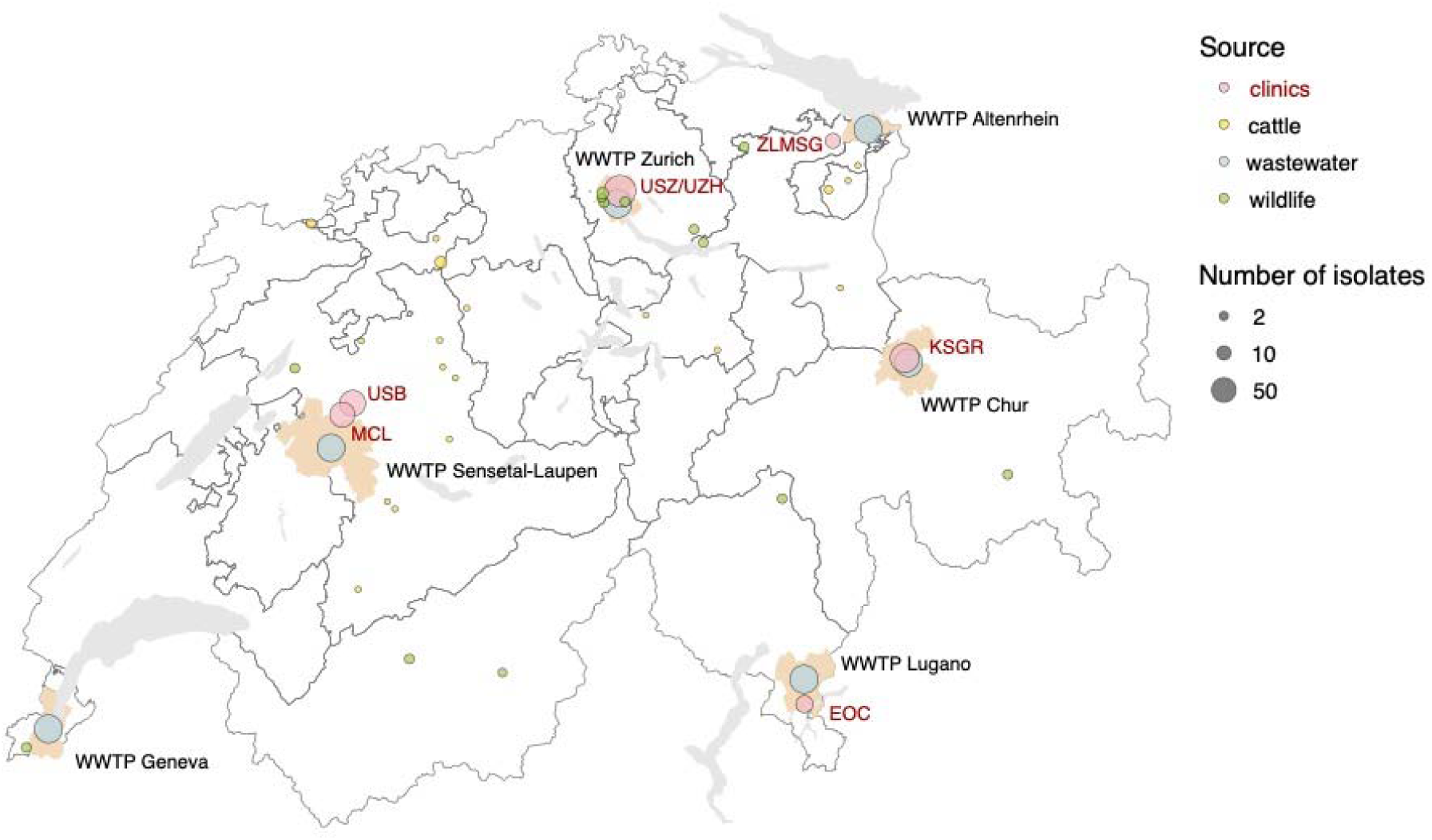
Geographic distribution of sampled ESBL-producing *E. coli* isolates across Switzerland. Map of Switzerland showing the distribution of sampled ESBL-*E. coli* isolates, with the size of the circle indicating the number of isolates and the colour representing the source. The sources include clinics (red), cattle (yellow), wastewater (blue), and wildlife (green). The locations of six wastewater treatment plants (WWTPs) from which ESBL-*E. coli* are isolated are marked by their corresponding catchments: WWTP Zurich, WWTP Altenrhein, WWTP Chur, WWTP Lugano, WWTP Sensetal-Laupen, and WWTP Geneva. Clinical settings from where the isolates were collected are labelled in red: Ente Ospedaliero Cantonale (EOC), Kantonsspital Graubünden (KSGR), Medizinische Laboratorien Niederwangen (MCL), Inselspital Bern (ISB), University Hospital Zurich (USZ/UZH), and Zentrum für Labormedizin (ZLMSG). The map was generated in R (v4.1.1) and modified in Inkscape (v1.1.1).

**Table 1:**
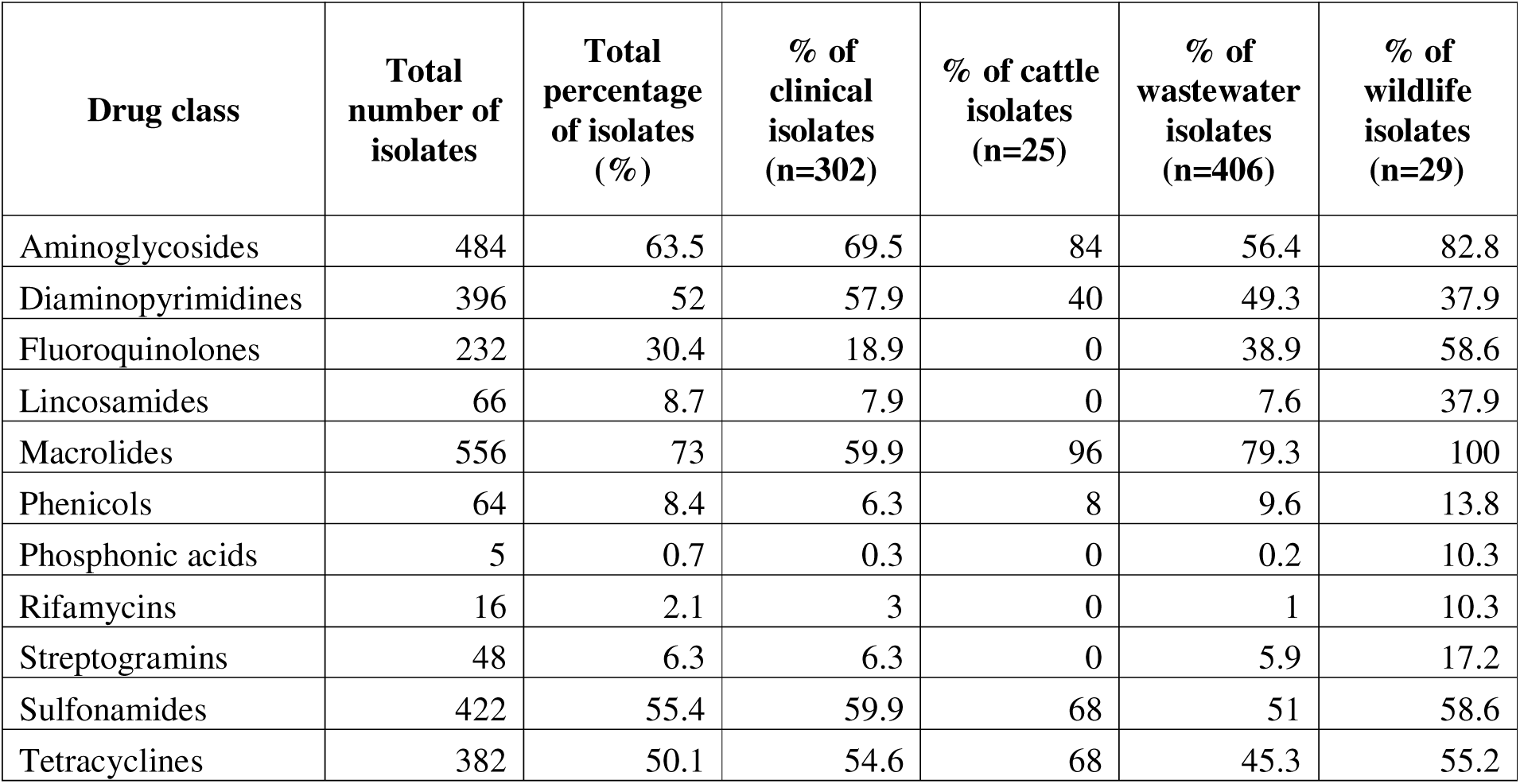
The table shows the number and percentage of isolates carrying resistance genes for each antibiotic class, with percentages also stratified by source (clinics, cattle, wastewater, and wildlife).

| Drug class | Total number of isolates | Total percentage of isolates (%) | % of clinical isolates (n=302) | % of cattle isolates (n=25) | % of wastewater isolates (n=406) | % of wildlife isolates (n=29) |
| --- | --- | --- | --- | --- | --- | --- |
| Aminoglycosides | 484 | 63.5 | 69.5 | 84 | 56.4 | 82.8 |
| Diaminopyrimidines | 396 | 52 | 57.9 | 40 | 49.3 | 37.9 |
| Fluoroquinolones | 232 | 30.4 | 18.9 | 0 | 38.9 | 58.6 |
| Lincosamides | 66 | 8.7 | 7.9 | 0 | 7.6 | 37.9 |
| Macrolides | 556 | 73 | 59.9 | 96 | 79.3 | 100 |
| Phenicols | 64 | 8.4 | 6.3 | 8 | 9.6 | 13.8 |
| Phosphonic acids | 5 | 0.7 | 0.3 | 0 | 0.2 | 10.3 |
| Rifamycins | 16 | 2.1 | 3 | 0 | 1 | 10.3 |
| Streptogramins | 48 | 6.3 | 6.3 | 0 | 5.9 | 17.2 |
| Sulfonamides | 422 | 55.4 | 59.9 | 68 | 51 | 58.6 |
| Tetracyclines | 382 | 50.1 | 54.6 | 68 | 45.3 | 55.2 |

### Diversity and phylogenetic distribution of ESBL-*E. coli*

A maximum-likelihood phylogenetic tree based on the core genome of 762 ESBL-producing *E. coli* isolates reveals clustering into two main clades (**Fig. 2**). EzClermont typing identified eight phylogroups: B2 (n=279, 37%), A (n=162, 21%), D (n=153, 20%), B1 (n=92, 12%), F (n=48, 6%), G (n=13, 2%), E (n=10, 1%), and cryptic (n=4, 0.5%) (**Table S4**). Additionally, one isolate (n=1, 0.1%) was categorized as unknown. Phylogroups A, B1, D, E, and F were present across all compartments, while B2 was predominantly detected in clinical samples (n=169, 61%) and wastewater (n=107, 38%), with three isolates (1%) from a single black-headed gull and none from cattle.

**Fig. 2:**
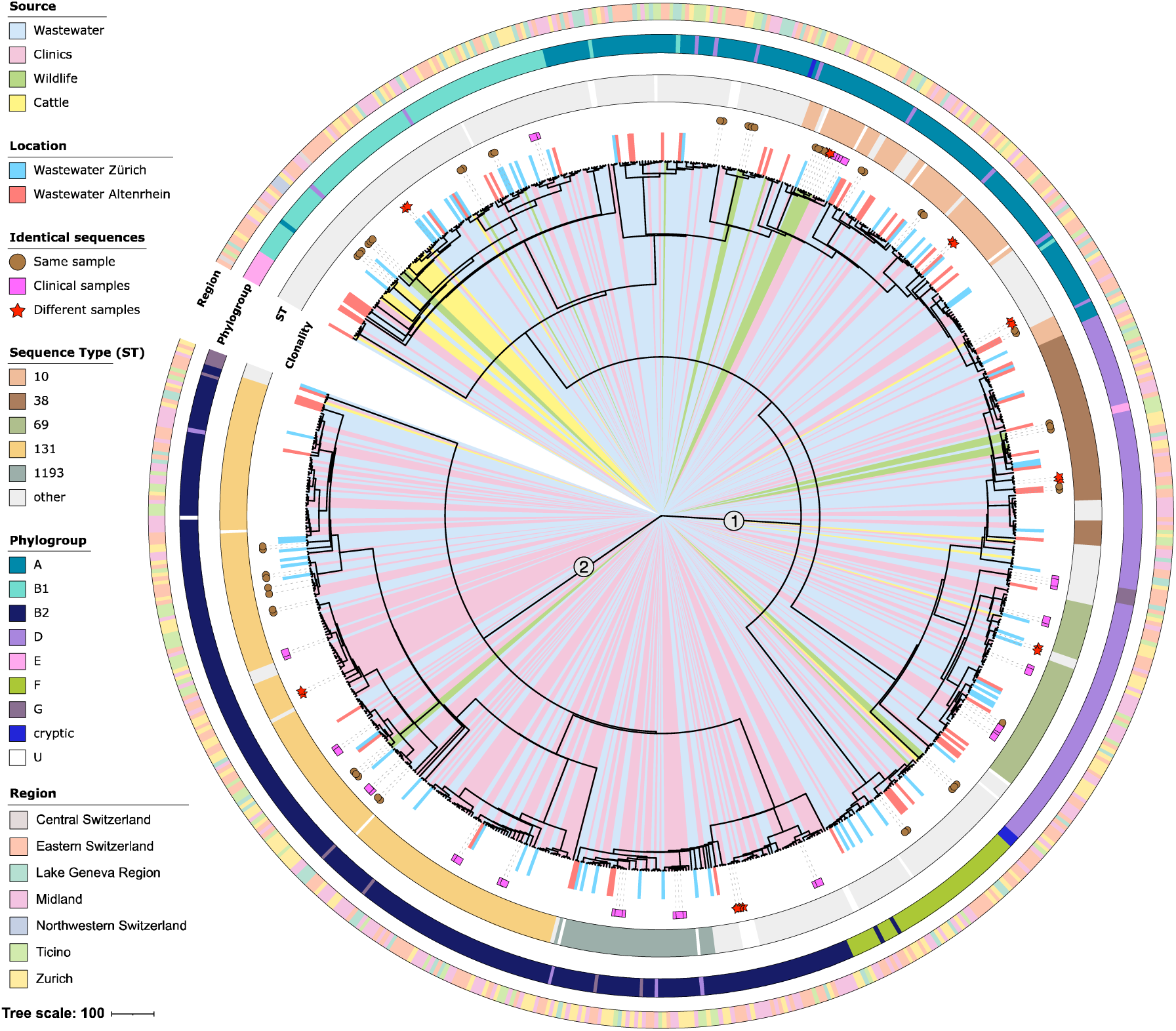
Phylogenetic structure and source distribution of ESBL-*E. coli* isolates. Maximum-likelihood phylogenetic tree of the core genome of 762 ESBL-*E. coli* isolates, illustrating phylogroups, sequence types (STs), collection sources, geographic regions, and clonal relationships. Each leaf corresponds to a distinct isolate, with source indicated by the inner colour band extending from the tree centre to the leaf label: wastewater (light blue), clinical settings (pink), cattle (yellow), and wildlife (green). Wastewater isolates from Zurich and Altenrhein treatment plants are specifically annotated in blue and red, respectively. Identical isolates are marked in the “Clonality” ring as retrieved from the same sample (brown circles), multiple clinical samples (pink squares), or different sources (red stars). Major STs (≥5% prevalence: ST38, ST69, ST131, ST10, ST1193) are highlighted with specific colours, while less frequent STs are shown in grey and undefined STs in white. Phylogroups (A, B1, B2, D, E, F, G, cryptic, and unknown) are represented by distinct coloured rings encircling the tree. The outer ring indicates the geographic region where each isolate was collected.

Among the isolates, 148 sequence types (STs) were identified (**Table S3**). The most prevalent was ST131 (n=197, 26%), followed by ST10 (n=66, 9%), ST38 (n=55, 7%), ST69 (n=53, 7%), and ST1193 (n=44, 6%) (**Fig. 2, Table S5**). Clinical samples were dominated by phylogroup B2 and ST131. In contrast, cattle-derived isolates (n=25) largely belonged to less common STs, with only one identified as ST69. Notably, ST131 isolates (n=197) were detected in humans, wastewater, and wildlife, but were absent in cattle.

The phylogenetic tree reveals extensive intermixing of ESBL-producing *E. coli* isolates across sources (clinics, wastewater, cattle, and wildlife) and regions (**Fig. 2**). Clade 1 is the most diverse and includes isolates primarily from phylogroups A, B1, and D, as well as less prevalent sequence types (ST10, ST38, and ST69). Clade 2 is dominated by ST131 and ST1193 isolates, with a marked concentration of clinical isolates. Notably, isolates from wastewater are widely dispersed in the tree. This holds for isolates from WWTPs with (e.g., Zurich) and without hospitals (e.g., Altenrhein) in their catchment areas, reflecting their genetic diversity independently from the hospital wastewater input. Wildlife and cattle isolates are similarly interspersed, with cattle isolates predominantly clustering within phylogroup B1. No exclusive clades were formed by isolates from a single source or region.

### Antibiotic resistant genes (ARGs) associated with ESBL-*E. coli*

A total of different 46 β-lactamase genes were detected among the ESBL-*E. coli* isolates (**Fig. 3A**, **Table S6**). In addition to *bla_CTX-M_*genes, other β-lactamase gene families such as *bla_TEM_*, *bla_OXA_*, and *bla_SHV_*were also identified, with some gene families showing frequent co-occurrence within individual isolates (**Extended Data** Fig. 1). *bla_CTX-M-15_* was the most prevalent ESBL gene, detected in 61% of wastewater, 57% of clinical, 38% of wildlife, and 12% of cattle isolates, followed by *bla_CTX-M-1_*, which was common in cattle (24%), present in wastewater (6%) and clinics (5%), but absent in wildlife. Additionally, 14 other *bla_CTX-M_* type genes were identified in all sources except cattle. The most prevalent were *bla_CTX-M-14_*, found in 21% of wildlife, 6% of wastewater, and 4% of clinical isolates, and *bla_CTX-M-55_*, which was present across wildlife (21%), wastewater (3%), and clinical sources (3%).

**Fig. 3:**
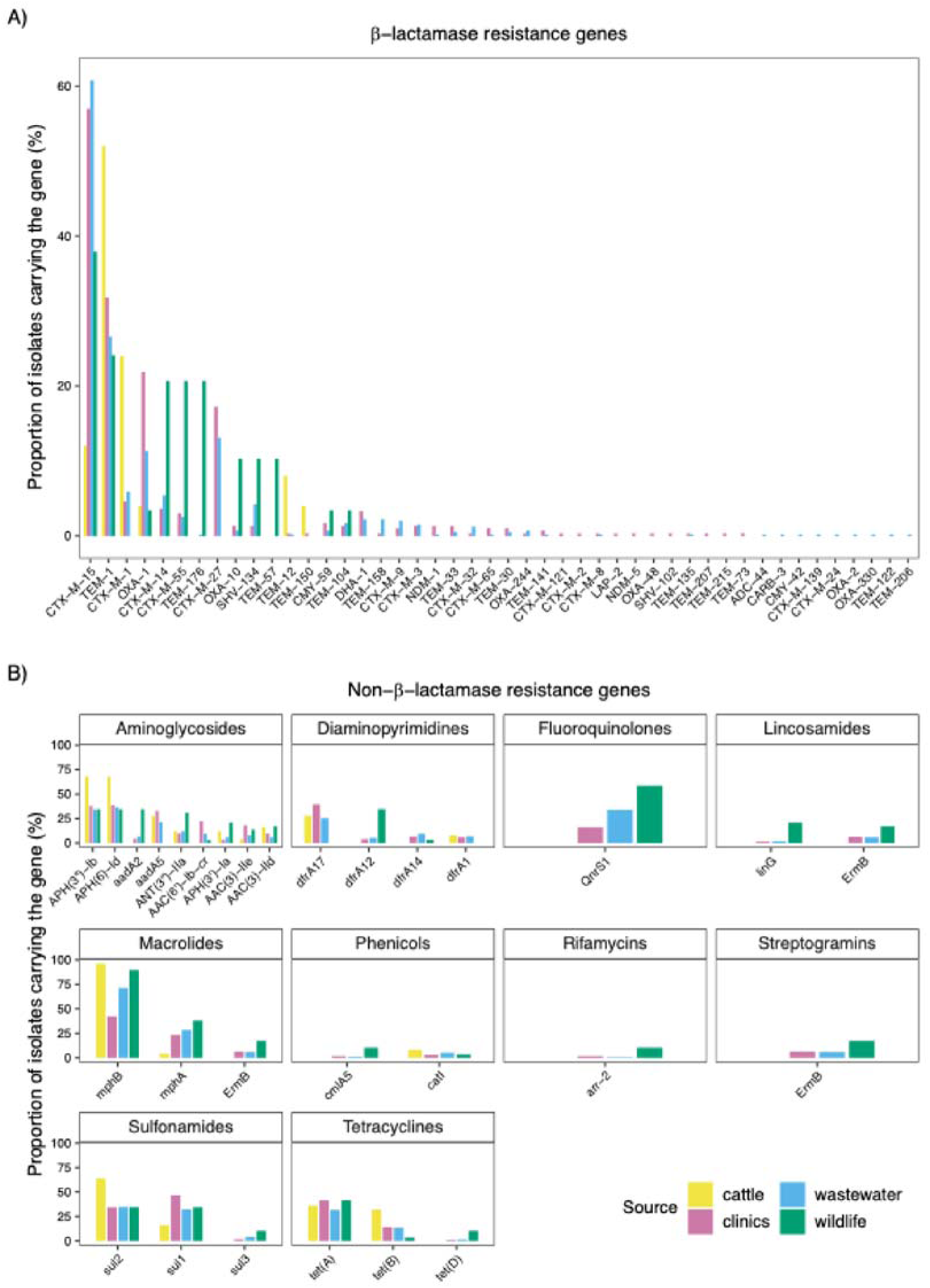
Distribution of β-lactamase and other antibiotic resistance genes across sources. (A) Proportion of isolates from clinics (pink), cattle (gold), wildlife (green), and wastewater (blue) carrying specific β-lactamase genes, identified using the Comprehensive Antibiotic Resistance Database (CARD). Each bar represents the proportion of isolates within each source harbouring the corresponding gene. (B) Proportion of isolates carrying resistance genes from other antibiotic classes, identified using CARD. Only genes present in ≥5% of isolates in at least one source are shown. Antibiotic classes include aminoglycosides, diaminopyrimidines, fluoroquinolones, lincosamides, macrolides, phenicols, phosphonic acids, rifamycins, sulfonamides, tetracyclines, and streptogramins. *ermB* is displayed in the lincosamides, streptogramins, and macrolides panels because it was annotated as conferring resistance to all three antibiotic classes. For an expanded CARD analysis, see **Fig. S4**.

*bla_TEM-1_* was the dominant *bla_TEM_* type gene, most frequently found in cattle (52%). *bla_SHV_* type genes were rare, with only *bla_SHV-102_* found exclusively in clinical isolates and *bla_SHV-134_* detected in clinical, wastewater, and wildlife isolates. *bla_OXA-1_* was the most prevalent *bla_OXA_* type gene, detected in all sources but mainly in clinical (22%) and wastewater (11%) isolates. *bla_OXA-10_* was predominantly found in wildlife (10%, n=3), but also in clinics (1%, n=4) and wastewater (1%, n=3). *bla_OXA-2_*, *bla_OXA-244_*, and *bla_OXA-48_* were found at low prevalence (<1%) in clinics and wastewater. Carbapenemase genes NDM-1 and NDM-5 were detected at low prevalence (<1%), while CARB-3 was identified in a single wastewater isolate.

Resistance genes were detected across 11 antibiotic classes beyond β-lactams, based on annotations from the Comprehensive Antibiotic Resistance Database (CARD, accessed November 4, 2024). These included macrolides (73% of isolates), aminoglycosides (64%), sulfonamides (55%), diaminopyrimidines (52%), tetracyclines (50%), fluoroquinolones (30%), lincosamides (9%), phenicols (8%), streptogramins (6%), rifamycins (2%), and phosphonic acids (0.7%), with notable variation across sources (**Fig. 3B**, **Table 1**). Macrolide resistance genes were present in 100% of wildlife isolates, 96% of cattle isolates, 79% of wastewater isolates, and 60% of clinical isolates. Similarly, aminoglycoside resistance genes were more frequent in cattle (84%) and wildlife (83%) than in clinics (70%) and wastewater (56%). Fluoroquinolones, lincosamides, phosphonic acids, streptogramins, and rifamycin resistance genes were absent in cattle but detected at varying levels in other sources. Fluoroquinolone resistance genes were present in 59% of wildlife, 39% of wastewater, and 19% of clinical isolates. Lincosamides resistance genes were most common in wildlife (38%) but rare in clinics (8%) and wastewater (8%). Streptogramin genes followed a similar pattern, with 17% in wildlife and 6% in both clinics and wastewater, while rifamycin resistance genes were found only in a small proportion of wildlife (10%), clinical (3%) and wastewater (1%) isolates. Wildlife had the highest prevalence of phosphonic acid resistance genes (10%), though they were also detected at low levels in clinical (0.3%) and wastewater (0.2%) isolates. Seventy-six percent (76%) of isolates were multidrug-resistant, carrying resistance genes to at least three antibiotic classes. This analysis considered resistance genes across the 11 non-β-lactam classes listed above (macrolides, aminoglycosides, sulfonamides, diaminopyrimidines, tetracyclines, fluoroquinolones, lincosamides, phenicols, streptogramins, rifamycins, and phosphonic acids), as well as β-lactamases. Notably, four isolates exhibited resistance genes to up to nine classes (**Extended Data Table 2**). The number of ARGs per isolate was similar across clinics, cattle, wastewater, and wildlife in most Swiss regions, with no significant differences detected in Eastern Switzerland, Lake Geneva Region, or Ticino (Kruskal-Wallis, p > 0.3). Marginally significant differences were observed in Zurich (p = 0.03) and Midland (p = 0.04), but no pairwise comparison remained significant after global Bonferroni correction (**Table S7**, **Extended Data** Fig. 2).

### Genetic similarity

Across all SNP thresholds, we observed 5,272 within-compartment and 5,318 between-compartment genetically similar pairs at 21-100 SNPs, 2,050 within-compartment and 1,841 between-compartment pairs at 1-20 SNPs, and 39 within-compartment and 2 between-compartment pairs at 0 SNPs (**Extended Data Table 3**). Within compartments, the proportion of genetically similar pairs calculated relative to all possible isolate pairs per compartment, to account for differences in sampling density, varied by source (**Fig. 4A**), with 7.7% of clinical isolates forming genetically similar pairs at 21-100 SNPs, compared to 2.3% in wastewater, 1.3% in cattle, and 2.9% in wildlife (**Extended Data Table 3**). The proportion of genetically similar pairs at stricter thresholds (1-20 and 0 SNPs) decreased across all compartments, with 5 zero-SNP clonal pairs in wastewater, 1 in wildlife, 1 in cattle and 32 in clinics. Between compartments, the highest proportion of genetically similar pairs was observed between clinics and wastewater (4.3% at 21-100 SNPs), followed by clinics-wildlife (1.9%), wastewater-wildlife (1.4%), and cattle-wastewater (0.4%) (**Fig. 4B**). Genetically identical isolate pairs (0 SNPs) were detected both within and between compartments (**Table S8**). The majority were observed within the same compartment, with multiple matches in clinics, as well as in wastewater, wildlife, and cattle. Clonal pairs between compartments were rare, with only two cases observed: one clinical isolate from St. Gallen that was detected in the wastewater of the nearest treatment plant (Altenrhein) and one from a Zurich hospital found in Zurich wastewater.

**Fig. 4:**
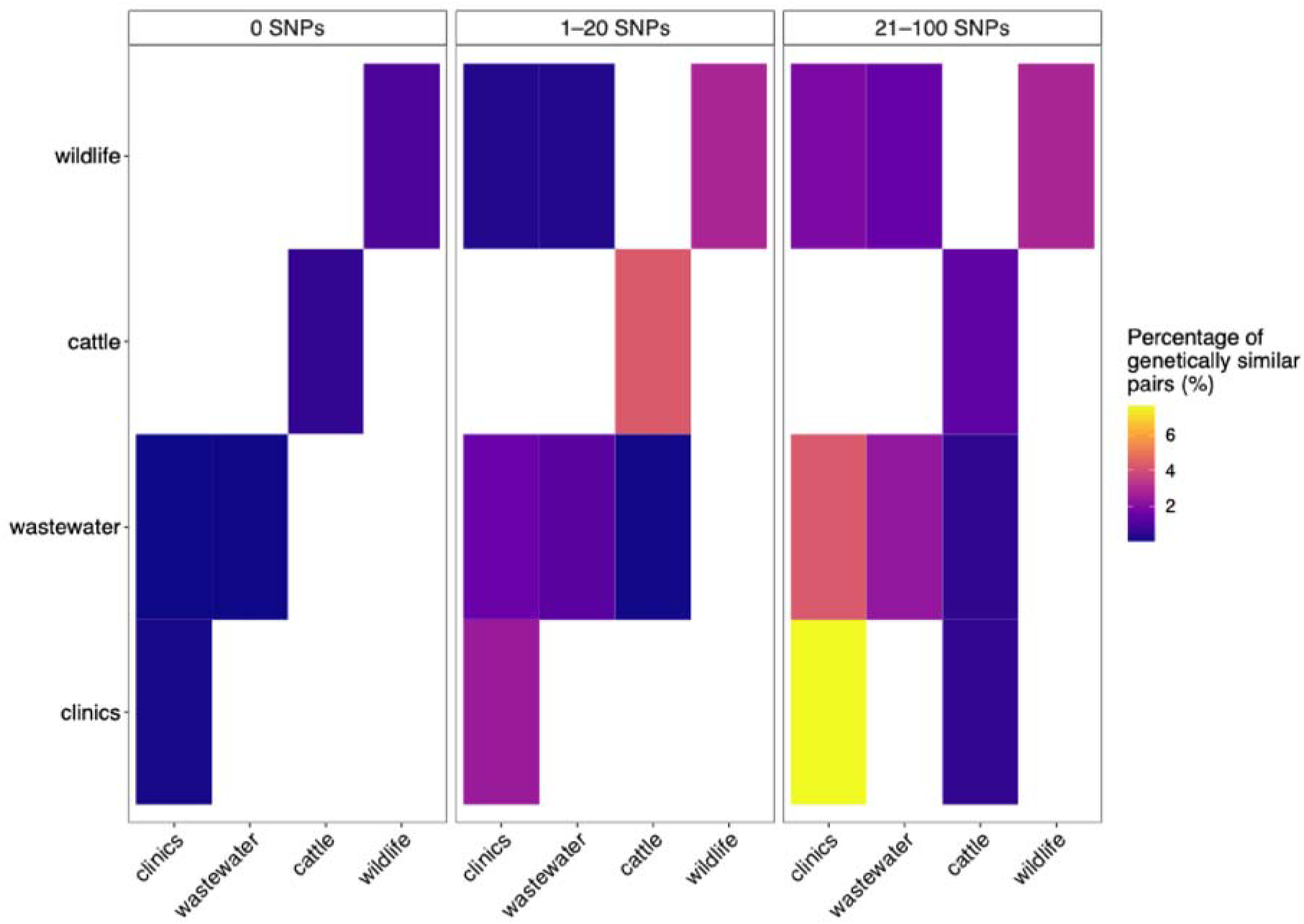
Within- and between-compartment genetic similarity. Percentage of genetically similar ESBL-*Escherichia coli* isolate pairs within and between compartments across SNPs thresholds. Diagonal cells represent within-compartment percentages; off-diagonal cells represent between-compartment percentages. White tiles indicate unobserved or zero values.

### Ancestral state reconstruction and transition analysis

Ancestral state reconstruction on 1,000 bootstrap maximum likelihood (ML) phylogenetic trees using the parsimony criterium demonstrated that transitions between sources in the full phylogeny of 792 ESBL-producing *E. coli* were not uniformly distributed (**Fig. 5**). Z-score analysis was applied to compare observed transitions to the null distribution, based on the approximation that Z-scores follow a standard normal distribution. This approach was supported by the near-normal distribution of transition counts under randomization in the consensus phylogeny (**Fig. S2**). Using this framework, transitions between community (wastewater) and clinics, as well as from wildlife to clinics, occurred significantly less frequently than expected under the null model (Z < −1.96). Transitions between other source pairs on the full phylogeny fell within the range expected under random permutations, with the exception of wildlife to cattle, which exhibited a slight excess of transitions relative to the null model, exceeding the central 95% confidence interval in 292 out of 1,000 bootstrap trees (**Fig. 5**, **Table S9**). Transitions involving wildlife and cattle were based on fewer isolates, due to the smaller representation of these compartments in the phylogeny (**Table S1**).

**Fig. 5:**
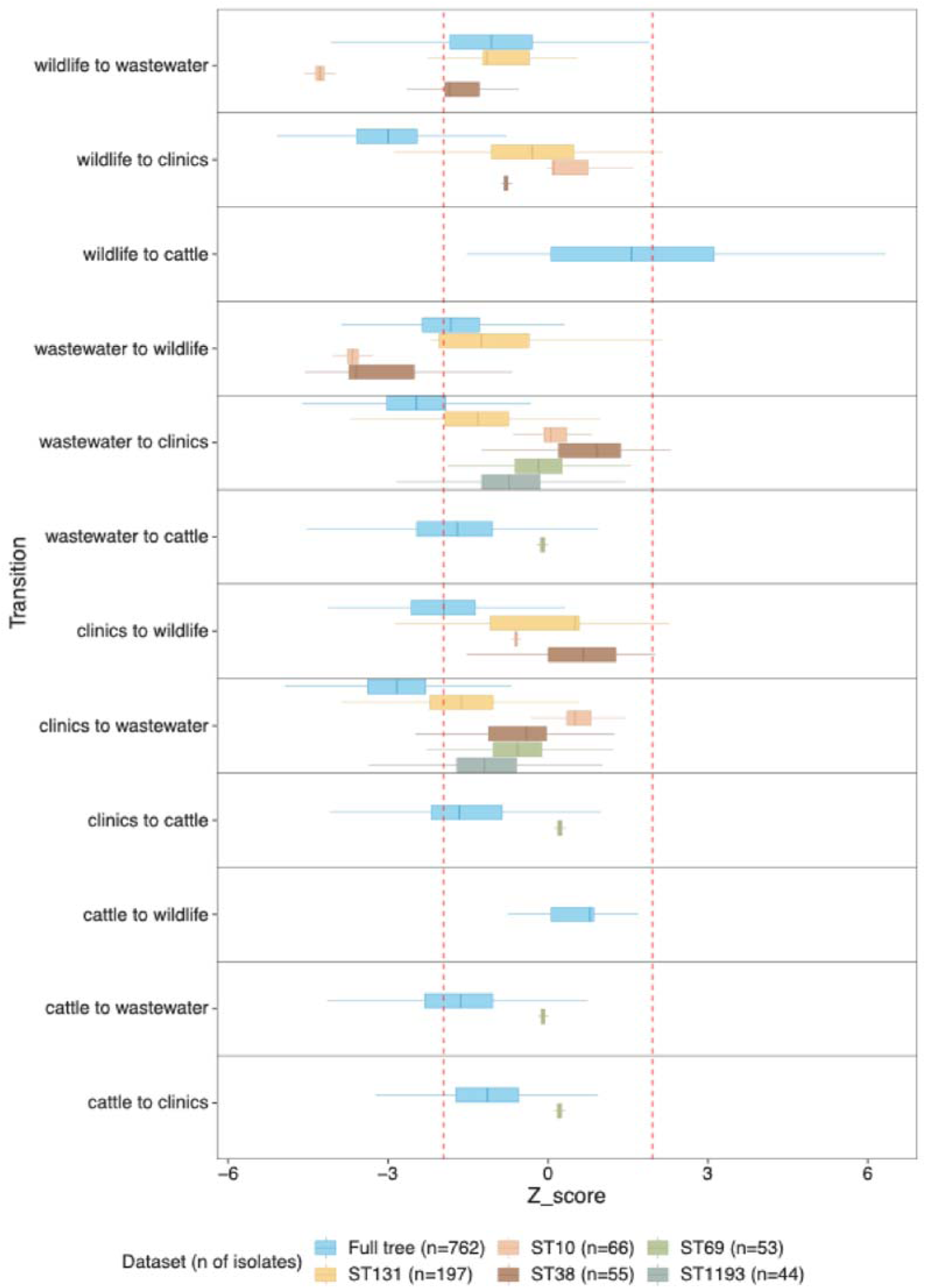
Z-score distributions of collection source transitions across 1,000 bootstrap phylogenies for the full dataset and dominant sequence types (STs). Transitions between collection sources (clinics, wastewater, wildlife, cattle) were inferred using maximum parsimony ancestral state reconstruction across 1,000 bootstrap maximum likelihood (ML) phylogenetic trees. For each tree, tip states were randomized 1,000 times preserving source frequencies to generate a null distribution of transition counts. Z-scores represent standardized deviations of observed transitions relative to the null. Each boxplot shows Z-score distributions per transition type, stratified by dataset subset (full dataset and the five dominant STs: ST131, ST10, ST38, ST69, ST1193). The legend shows the number of isolates retained in the analysis for each ST. Red dashed lines indicate ±1.96, corresponding to the central 95% confidence interval under the null model. Transitions were reconstructed using the *acctran* and *ancestral.pars* functions from the ape package (v5.7.1) in R (v4.1.2). Positive Z-scores indicate transitions occurring more frequently than expected by chance; negative Z-scores indicate less frequent transitions.

When analysed within the five most dominant sequence types (ST131, ST10, ST38, ST69, ST1193), the non-random patterns observed in the full dataset were not retained. Transitions between community (wastewater) and clinics occurred at frequencies consistent with random expectations in all five STs, with no significant deviations observed. In ST10, however, transitions between wildlife and community were significantly underrepresented in both directions (Z < −1.96). A similar pattern was observed in ST38, where transitions from community to wildlife occurred significantly less frequently than expected. Transitions among all other source pairs within individual STs, including between cattle, wildlife, clinics, and community, did not deviate from random expectations. The same pattern was observed when analyses were restricted to a subtree including only wastewater (n = 406) and clinical (n = 302) isolates, to control for potential sampling biases (**Extended Data** Fig. 3). On the full subset phylogeny, transitions between wastewater and clinics were consistently less frequent than expected under the null model, across all 1,000 bootstrap trees (**Table S13**). Within sequence types, however, transitions between these sources conformed to random expectations.

## DISCUSSION

Our study highlights that within clinically relevant human-associated sequence types (STs), ESBL-producing *E. coli* circulate between clinics and the broader community, with wastewater surveillance capturing strains that overlap with those found in clinical settings. Although the phylogeny showed extensive intermixing across sources and no source- or region-specific clades, genetic similarity was higher within than between compartments. Clonal isolates (0 SNPs) were rare between compartments (n = 2, both between clinics and wastewater), in contrast to 32 within-clinic and a few within other sources. Ancestral state reconstruction on the whole phylogeny revealed significantly fewer transition events than expected by chance between clinics and both wastewater (used as a proxy for community shedding) and wildlife, suggesting partial compartmentalization and limited spillover, and indicating potential epidemiological separation. The limited connectivity between wildlife and clinical settings may reflect reduced exposure opportunities between wildlife and the human-built environment in Switzerland. Conversely, transitions between livestock and wildlife exceeded the null expectation in a small subset of bootstrap trees, which may indicate some degree of exchange or reflect reduced power to detect significant structure due to small sample sizes (28). Similarly, the observation that transitions between wildlife or livestock and wastewater fell within the range expected under randomization may reflect the fact that wastewater is not entirely distinct from these sources, as it can receive input from agricultural runoff and surrounding environments, such as slaughterhouses.

When analysed by sequence type (ST), this structured signal diminished. ST131, the most prevalent lineage (26%), showed no deviation from random expectations in transitions between clinics and the community (as captured by wastewater), suggesting free circulation between symptomatic and asymptomatic hosts, and highlighting the potential of wastewater as an indicator of human gut colonization (23). ST131 dominated clinical isolates, but it was absent from cattle, aligning with studies reporting genetic divergence between human-and cattle-derived *E. coli* strains (24). A slight, non-significant excess of transitions from clinics to wildlife was also observed, suggesting rare spillover events. Supporting this, one black-headed gull isolate belonging to ST131 clustered with wastewater and clinical isolates, suggesting cross-compartment interactions. Wildlife may act as a reservoir for clinically relevant AMR, particularly in species interacting with WWTPs, consistent with evidence of human-wildlife AMR exchange in urban birds in Kenya and of gulls accumulating AMR from WWTP sediment basins in Sweden (25–28). Nonetheless, transmission from wildlife to clinics or wastewater appeared rare (11).

ST1193 and ST69, two other human-associated lineages (29, 30), also showed transition frequencies between clinics and community consistent with random expectations, supporting their capacity to circulate across these compartments resulting in their epidemiological success (30–32). These results suggests that the separation observed in the full tree is not due to universal barriers but rather reflects lineage-specific ecological behaviour. In contrast, the more ecologically versatile ST10 and ST38 (33, 34) exhibited reduced transitions between community and wildlife, indicating potential source-specific adaptation or reduced contact between wildlife and the human-built environment in Switzerland.

Genetically identical strains were rarely shared between compartments. In both observed zero-SNP matches between wastewater and clinical isolates, the wastewater isolate was detected two to three weeks after the clinical counterparts, suggesting that clinical cases may contribute to subsequent detection in wastewater through shedding (35) or that these clones are more broadly circulating within the community, including via asymptomatic carriage. The highest proportion of genetically similar between-compartments pairs occurred between clinics and wastewater (4.3% at 21–100 SNPs), suggesting that exchange occurs mostly through closely related but non-identical strains, likely due to indirect transmission, rapid divergence following environmental entry, or limited sampling resolution. These findings emphasize how SNP thresholds shape interpretations of genetic relatedness and underscore the need of context-specific thresholds in genomic surveillance (30).

β-lactamase genes were widely distributed across all sources, with 46 genes detected, consistent with global reports of widespread β-lactam resistance (36, 37). *bla_CTX-M-15_* was the most prevalent, detected in 61% of wastewater and 57% of clinical isolates, reinforcing its dominance in human-associated environments (38). Its abundance in both compartments suggests wastewater may reflect the broader community reservoir and detect resistance circulating before clinical detection. Supporting this, wastewater from the Altenrhein catchment (without hospital input) showed comparable ESBL-*E. coli* diversity to that in Zurich (with hospital input), highlighting the contribution of community sources and the value of wastewater monitoring for population-level AMR surveillance (39). In cattle, *bla_CTX-M-1_* was common, consistent with its prevalence in food-producing animals (40), while *bla_CTX-M-14_*, previously reported in cattle, was absent, indicating possible temporal shifts in gene prevalence. In wildlife, *bla_CTX-M-15_*, *bla_TEM_*, *bla_SHV_*, and *bla_OXA-10_*, were present, but *bla_CTX-M-1_* was not, mirroring regional differences observed in wild ruminants and boar in Germany (41, 42). These results point to wildlife as a reservoir for specific resistance genes and emphasize the need for cross-sectoral AMR monitoring in line with One Health principles (11, 43). Among the 76% of isolates in our dataset carrying resistance to at least three antibiotic classes, consistent with global reports estimating a 66% prevalence of multidrug resistance among ESBL-*E. coli* (31, 44–46), one duck-derived isolate exhibited genotypic resistance to nine antibiotic classes. This extreme genotype reinforces the role of wildlife in AMR dissemination and underscores growing global concerns about the environmental spread of resistance (47, 48).

Although we systematically collected wastewater isolates, cattle, wildlife, and clinical isolates were obtained through convenience sampling, potentially limiting representativeness and introducing source-specific biases. A subset of clinical isolates was restricted to invasive infections and did not include colonization, which may underestimate within-clinic diversity. The small sample sizes for wildlife and cattle may have underestimated AMR diversity and reduced statistical power to detect transitions involving these compartments. Furthermore, livestock sampling focused exclusively on cattle, omitting pigs and poultry, which may have closer epidemiological links to humans. Wildlife samples were primarily collected outside WWTP catchments, potentially overlooking interactions with peri-urban or slaughterhouse environments that may serve as transmission interfaces. Moreover, while we interpret ESBL-*E. coli* in wastewater as reflecting community shedding, wastewater may also include hospital, industrial, and animal inputs, and strains may persist in sewage networks independent of human input via mechanisms such as biofilm formation (49–51).

Our results indicate that while transitions between sources occur, compartments remain partially structured on the full phylogeny, particularly between clinics and wastewater. This separation disappears within clinically relevant human-associated sequence types (ST131, ST69, ST1193), where the absence of structure suggests dynamic circulation between clinical settings and the broader community. Notably, wastewater isolates captured lineages also detected in clinical settings, supporting its utility as a proxy for community-level AMR carriage. Although zero-SNP matches between compartments were rare, the predominance of genetically similar isolates within clinics and wastewater points to localized clonal expansion and potential indirect exchange. These findings highlight how the interpretation of compartmental connectivity depends on the analytical resolution used, and underscore the importance of context-specific SNP thresholds in One Health genomic surveillance (52). While expanding surveillance in underrepresented reservoirs such as wildlife and cattle remains important, our study reinforces the value of wastewater monitoring for capturing clinically relevant ESBL-*E. coli* circulating in the human population and supports its integration into national One Health surveillance frameworks.

## ONLINE METHODS

### Sample collection

A total of 834 ESBL-*E. coli* isolates were collected from wastewater (n = 438), healthcare facilities (n = 328), cattle (n = 26) and wildlife (n = 42) across Switzerland from 2021 to 2023 (**Fig. 1**, **Extended Data Table 1**). Cattle and wildlife isolates were obtained through two independent field studies conducted during the same period, which focused on antimicrobial resistance in Swiss dairy herds and free-roaming wild animals, including mammals and birds (53, 54), and correspond to the same isolates processed and identified in a previous study (55). Wastewater samples were obtained from six wastewater treatment plants (WWTPs) using 24-hour flow-proportional composite sampling. Samples were transported to the laboratory on ice, stored at 4°C, and processed within 48 hours. To isolate ESBL-*E. coli*, 100 µL of undiluted wastewater was plated onto CHROMagar Orientation supplemented with ESBL selective supplement (CHROMagar, France). Presumptive ESBL-*E. coli* colonies were identified by their characteristic dark pink to reddish coloration. Every four weeks, three to eight colonies per WWTP were streaked on LB agar (Merck KGaA, Germany), incubated at 37°C for 20–24 h, and further cultured in Luria Broth (Merck KGaA, Germany). Single-colony cultures were preserved in 12% glycerol stocks at −70°C for downstream analyses. Clinical isolates were retrospectively collected from six healthcare facilities across Switzerland: Ente Ospedaliero Cantonale (Ticino), Kantonsspital Graubünden (Graubünden), MCL Medizinische Laboratorien Niederwangen (Bern), Inselspital (Bern), University Hospital Zurich (Zurich), and Zentrum für Labormedizin (St. Gallen). All clinical isolates were confirmed ESBL-producing *E. coli* as part of routine diagnostic workflows and were selected accordingly for this study. In facilities where the number of available ESBL-*E. coli* isolates exceeded sequencing capacity (e.g., Zurich, Bern, and Chur), a random subset was selected. Isolates originated from individuals screened for ESBL resistance were revived from 12% glycerol stocks or Microbank Cryovial Storage Systems (Pro-Lab Diagnostic, Canada), streaked onto LB agar, incubated at 37°C for 20–24 hours, and cultured in Luria Broth for DNA extraction.

### DNA Extraction and whole-genome sequencing

DNA was extracted using the DNeasy Blood and Tissue kit (Qiagen, Hilden, Germany) following the manufacturer’s instructions. Whole-genome sequencing (WGS) was performed using the LITE Library Prep method on an Illumina NextSeq 1000 P2 flow cell (300 bp paired-end) at the Earlham Institute, UK (56). Sequencing was conducted in two batches (June 2023 and January 2024) for 834 ESBL-producing *E. coli* isolates. Unique Dual Indexes were used to minimize index hopping (57). Quality control measures included duplicate samples, negative extraction controls (AE blanks), and reference isolates of E. coli (strain 1 and 2) and Klebsiella pneumoniae (strain 1 and 2). Negative controls (n = 12; 8 in batch 1, 4 in batch 2) were processed alongside study samples to monitor for contamination or sequencing artefacts. Reference *E. coli* strains (strain 1: n = 8 in batch 1, n = 1 in batch 2; strain 2: n = 8 in batch 1, n = 1 in batch 2) had been previously sequenced using alternative platforms and were included to confirm sequencing consistency. *K. pneumoniae* strains (strain 1: n = 8 in batch 1, n = 1 in batch 2; strain 2: n = 5 in batch 1, n = 2 in batch 2) were used to verify that the sequencing and bioinformatics pipelines could accurately distinguish *E. coli* from other species.

### Bioinformatic analyses and isolates characterization

The bioinformatic workflow is summarized in **Fig. S3**. Illumina reads were trimmed with trimmomatic v 0.35 in paired-end mode, removing Illumina Nextera adapters with specific trimming parameters (2:30:10:1) and applying quality-based trimming with a 4bp sliding window and an average quality threshold of 20 (58). Reads shorter than 36 bp were removed. Read quality before and after trimming was assessed using FastQC v0.11.4 (59). Screening for contamination was performed using FastQ Screen v0.15.3 (60), which maps reads against a panel of reference genomes to detect potential co-isolated or misidentified species. To this end, we downloaded seven reference genomes from NCBI corresponding to bacterial species known to grow on CHROMagar ESBL plates and potentially co-occur with *E. coli*: *Acinetobacter baumannii* (ASM863263v1), *Citrobacter freundii* (ASM381234v1), *Escherichia coli* (MG1655), *Enterobacter roggenkampii* (ASM172980v1), *Klebsiella pneumoniae* (ASM24018v2), *Pseudomonas aeruginosa* (ASM676v1), and *Proteus mirabilis* (ASM6996v1). Filtered reads were de novo assembled using SPAdes v3.15.9, without further error correction and set to ’careful’ (61). Assembly quality was assessed with QUAST v5.2.0 (**Table S2**) (62). Genome annotation was performed using Bakta v1.9.1, specifying *E. coli* as the genus and species (63). Sequence types (STs) were assigned using MLST v.2.23.0 with the Achtman scheme (https://github.com/tseemann/mlst). Phylogenetic groups were determined using EzClermont v.0.6.3 based on an *in-silico* PCR approach (64). Serotyping was performed using ecoli-serotyping (https://github.com/phac-nml/ecoliserotyping).

Antibiotic resistant genes (ARGs) were identified using Abricate v1.0.1 (65), blasting against the Comprehensive Antibiotic Resistance Database (CARD, accessed 4th November 2023) (66). ARGs were classified in antibiotic classes based on CARD annotations. Genes conferring resistance to at least four different classes and identified as efflux pumps were classified separately. Efflux pumps and genes intrinsic to *E. coli* were excluded from the main analyses but included in the supplementary material (**Fig. S4**). Statistical differences in the prevalence of ARGs among sources and regions were assessed using a Kruskal-Wallis and Dunn’s pairwise test with global Bonferroni adjustment. Regions are displayed in **Fig. S1**.

### Phylogenetic reconstruction and genetic similarity

A pangenome was defined using Roary v3.13.0, identifying a total of 29,477 gene clusters, of which 2,312 core genes (present in ≥ 99% of isolates) were retained for phylogenetic analysis. The core genome was aligned using MAFFT (67) and concatenated into a single alignment comprising 2,099,419 nucleotide positions. Recombinant regions were identified and masked using ClonalFrameML v1.13 (68). A maximum-likelihood phylogeny was constructed using IQ-TREE with automatic model selection and rooted with an *Escherichia albertii* reference genome (NCBI Accession: ASM1690475v2) included in the pangenome analysis (69). The resulting tree was annotated and visualized using iTOL (70). Pairwise SNP differences between isolates from the same or different compartments (clinics, cattle, wastewater, and wildlife) were computed using snp-dists v0.8.2 on the recombination-free core genome alignment (https://github.com/tseemann/snp-dists). Identical sequences from the same sample were collapsed post-SNP calculation, retaining one representative per sample to avoid inflation of pairwise comparisons in the analysis of genetically similar pairs (**Table S11**). The proportion of genetically similar pairs was calculated in R (v4.3.1), within and between compartments, using SNP thresholds of 0, 1-20, and 21-100 SNPs to assess genetic similarity and potential transmission links.

### Ancestral state reconstruction and transition analysis

Ancestral state reconstruction of isolate collection sources (clinics, wastewater, cattle, wildlife) was conducted using maximum parsimony to infer the most likely source transitions along phylogenetic tree branches (71). Wastewater was used as a proxy for community-level ESBL-*E. coli*, as it captures bacteria shed by the general population. Tip states were assigned based on isolate origin and mapped onto both the consensus maximum-likelihood (ML) phylogeny and 1,000 bootstrap ML trees. Ancestral states at internal nodes were inferred using the *acctran* and *ancestral*.*pars* functions in the ape package (v5.7.1) within R (v4.1.2) (72). Transitions between sources were quantified for each phylogeny by identifying changes in reconstructed states along branches, excluding self-transitions (i.e., source remaining unchanged). To evaluate whether observed transitions deviated from random expectations, a null model was implemented whereby tip states were permuted 1,000 times for each bootstrap tree, maintaining the original frequency distribution of sources. For each randomized replicate, ancestral states were re-estimated using maximum parsimony, and the number of transitions between each pair of sources was recorded, resulting in a null distribution of transition counts for every source combination. For each randomization, ancestral states were reconstructed and transitions were counted, generating a null distribution for each source pair. For each source transition, a Z-score was calculated according to the formula:

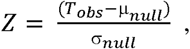

where *T_obs_* is the observed transition count, and μ_null_ and σ_null_ are the mean and standard deviation of the null distribution, respectively. Positive Z-scores indicated transitions more frequent than expected by chance, and negative Z-scores indicated fewer transitions. Z-scores exceeding ±1.96 were considered significant at the central 95% confidence level. Analyses were performed on the full dataset and separately on the five dominant sequence types (ST131, ST10, ST38, ST69, ST1193), by pruning the phylogenies to retain only isolates from each ST. An additional analysis was conducted using only isolates from wastewater and clinical sources and to the five dominant STs within this subset. Ancestral reconstruction, randomization testing, and Z-score computation were repeated accordingly.

## DATA AVAILABILITY

The sequence data is available at SRA under the BioProject ID PRJNA1271740. The bioinformatic pipeline, implemented using the Snakemake workflow management system is available at https://github.com/sheenaconforti/ecoli-wgs/. All scripts used for data analysis and figure generation, are available at https://github.com/EawagPHH/wgs_esblecoli.

## FUNDING STATEMENT

The work was funded through the Swiss National Science Foundation grants 192763 to TRJ, and 205933 to TS, and through a Swiss Federal Office of Public Health grant to Christoph Ort and TRJ.

## ACKNOWLEDGEMENTS

We thank Lea Caduff, Charlie Gan, Laura Brülisauer, Camille Hablützel, and Aurélie Holschneider for their assistance in processing wastewater samples, Gladys Martinetti Lucchini for providing clinical isolates, and Aitana Neves for providing feedback. We are grateful to the Wastewater-based Surveillance for Infectious Diseases and Surveillance (WISE) group for their valuable insights and contributions to the general discussion. We also extend our appreciation to Timothy G. Vaughan, Etthel M. Windels, and Cecilia Valenzuela Agüí for their support with the phylodynamic analyses. Finally, we thank the employees of the wastewater treatment plants IDA CDA Lugano (Ticino), ARA Werdhölzli (Zurich), ARA Chur (Graubünden), ARA Sensetal Laupen (Bern), and STEP d’Aïre Genève (Geneva) for providing wastewater samples.

## EXTENDED DATA TABLES

**Extended Data Table 1:** Summary of the number of isolates collected from various sources, including wastewater, clinics, wildlife, and cattle, along with their respective collection timeframes and locations. The total number of isolates collected from each source category is indicated in parentheses next to the source name.

| Source | Location | Number of isolates | Collection timeframe |
| --- | --- | --- | --- |
| Wastewater<br>(n=438) | ARA Altenrhein | 73 | 16.11.2021 - 21.11.2022<br>(in addition 5 samples per location from 30.10.2023) |
|  | ARA Chur | 72 |  |
|  | ARA Sensetal Laupen | 71 |  |
|  | ARA Werdhölzli<br>Zürich | 74 |  |
|  | IDA CDA Lugano | 74 |  |
|  | STEP d'Aïre Genève | 74 |  |
| Clinics<br>(n=328) | EOC | 19 | 06.01.2021 - 21.12.2022 |
|  | KSGR | 78 | 01.11.2021 - 11.12.2022<br>(in addition 2 samples from 2019 and 2 samples from 2020) |
|  | MCL | 54 | 01.11.2021 - 08.12.2022 |
|  | USB | 61 | 03.11.2021 - 28.10.2022 |
|  | USZ | 103 | 01.02.2022 - 01.01.2023 |
|  | ZLMSG | 13 | 08.05.2022 - 20.12.2022 |
| Wildlife<br>(n=42) | AUS (Near Altenrhein) | 3 | 5.12.2022 - 01.06.2023 |
|  | Bern | 3 |  |
|  | Geneva | 3 |  |
|  | Graubünden | 3 |  |
|  | St. Gallen | 6 |  |
|  | Ticino | 3 |  |
|  | Wallis | 5 |  |
|  | Zurich | 16 |  |
| Livestock<br>(n=26) | Appenzell | 4 | 4.11.2021 - 14.02.2023 |
|  | Basel | 3 |  |
|  | Bern | 8 |  |
|  | Lucern | 1 |  |
|  | Schwyz | 2 |  |
|  | Solothurn | 7 |  |
|  | St. Gallen | 1 |  |

**Extended Data Table 2:**
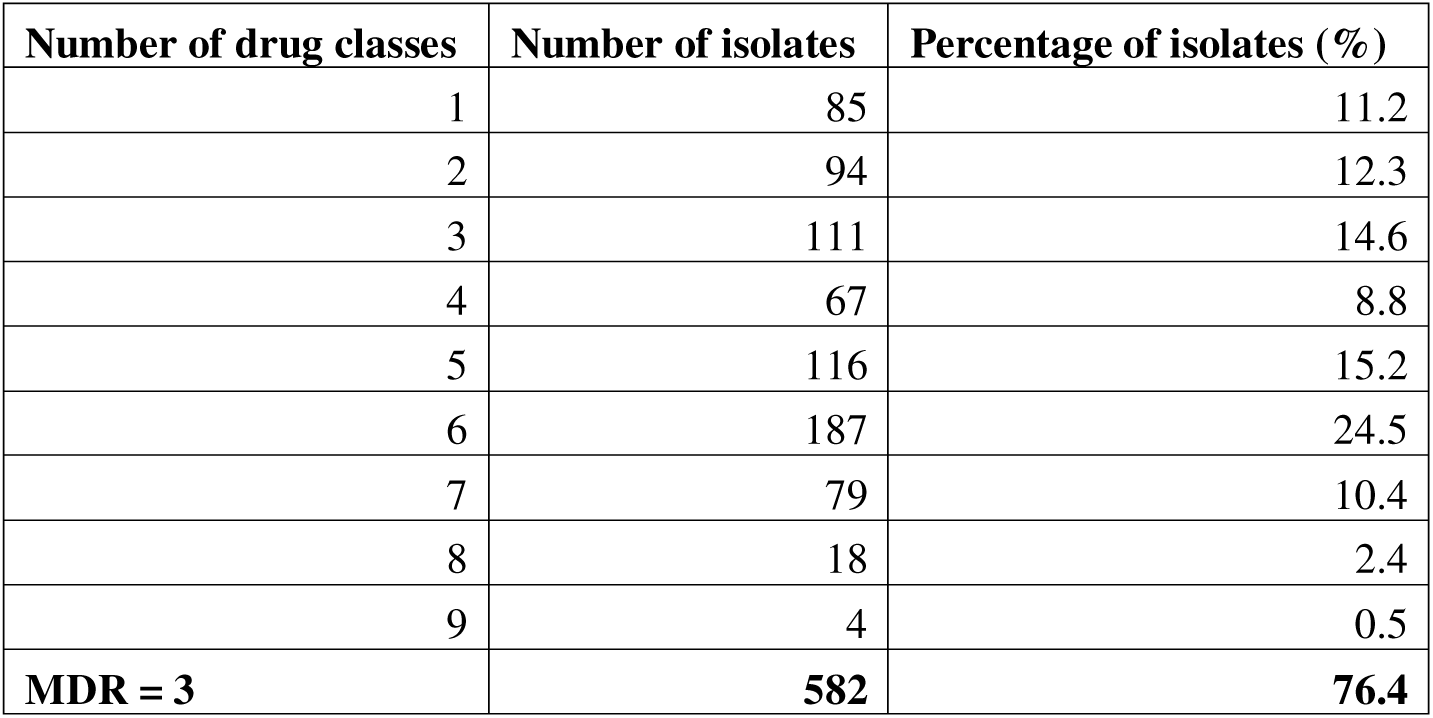
Distribution of isolates by number of drug resistance classes. The table shows the number and percentage of isolates resistant to varying numbers of drug classes, based on genes detected from the Comprehensive Antibiotic Resistance Database (CARD, accessed 4th November 2023). Multidrug resistance is abbreviated as “MDR”, and is defined as resistance to three or more classes. Classes considered were: aminoglycosides, β-lactamases, fluoroquinolones, diaminopyrimidines, macrolides, sulfonamides, tetracyclines, phenicols, phosphonic acids, lincosamides, and rifamycins.

**Extended Data Table 3:** Genetically similar *Escherichia coli* isolate pairs, including extended-spectrum β-lactamase (ESBL)-producing strains, detected within and between compartments across SNP thresholds (0, 1–20, and 21–100 SNPs). The upper section reports total pair counts by comparison type (within vs. between). The middle and lower sections show the percentage and count (n) of genetically similar pairs within and between compartments, respectively. Percentages are calculated relative to the total number of possible isolate pairs within or between the corresponding compartments. Analysis was performed in R (v4.1.1).

| Comparison | 21-100 SNPs | 1-20 SNPs | 0 SNPs |
| --- | --- | --- | --- |
| **Number of genetically similar pairs** |  |  |  |
| Between | 5318 | 1841 | 2 |
| Within | 5272 | 2050 | 39 |
| **Percentage of genetically similar pairs within compartments (%)** |  |  |  |
| cattle | 1.30 (n=3) | 4.33 (n=10) | 0.43 (n=1) |
| clinics | 7.69 (n=3497) | 2.50 (n=1138) | 0.07 (n=32) |
| wastewater | 2.33 (n=1769) | 1.19 (n=899) | 0.01 (n=5) |
| wildlife | 2.86 (n=3) | 2.86 (n=3) | 0.95 (n=1) |
| **Percentage of genetically similar pairs between compartments (%)** |  |  |  |
| clinics vs wastewater | 4.32 (n=5087) | 1.54 (n=1816) | 0.0017 (n=2) |
| cattle vs wastewater | 0.38 (n=33) | 0.04 (n=3) | NA |
| clinics vs wildlife | 1.88 (n=85) | 0.22 (n=10) | NA |
| wastewater vs wildlife | 1.44 (n=84) | 0.21 (n=12) | NA |
| cattle vs clinics | 0.44 (n=29) | NA | NA |

## EXTENDED DATA FIGURES

**Extended Data Fig. 1:**
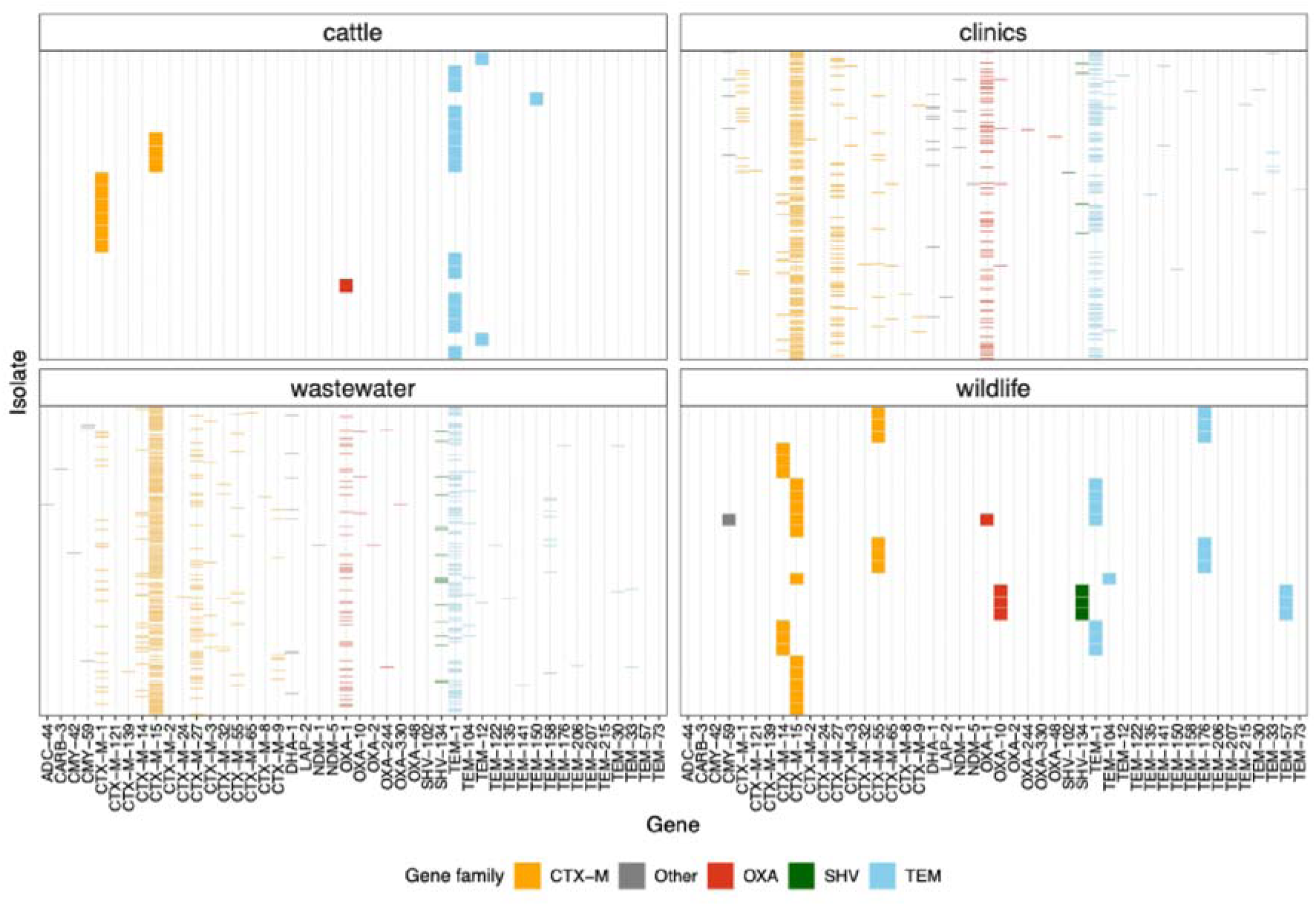
Co-occurrence of β-lactamase genes across ESBL-producing *Escherichia coli* isolates stratified by source (clinics, cattle, wastewater, and wildlife). The heatmap displays individual isolates (rows) and β-lactamase genes (columns). Each tile indicates the presence of a specific gene in a given isolate identified using the Comprehensive Antibiotic Resistance Database (CARD). Genes are grouped and color-coded by gene family: CTX-M (orange), TEM (skyblue), SHV (green), OXA (red), and Other (grey).

**Extended Data Fig. 2:**
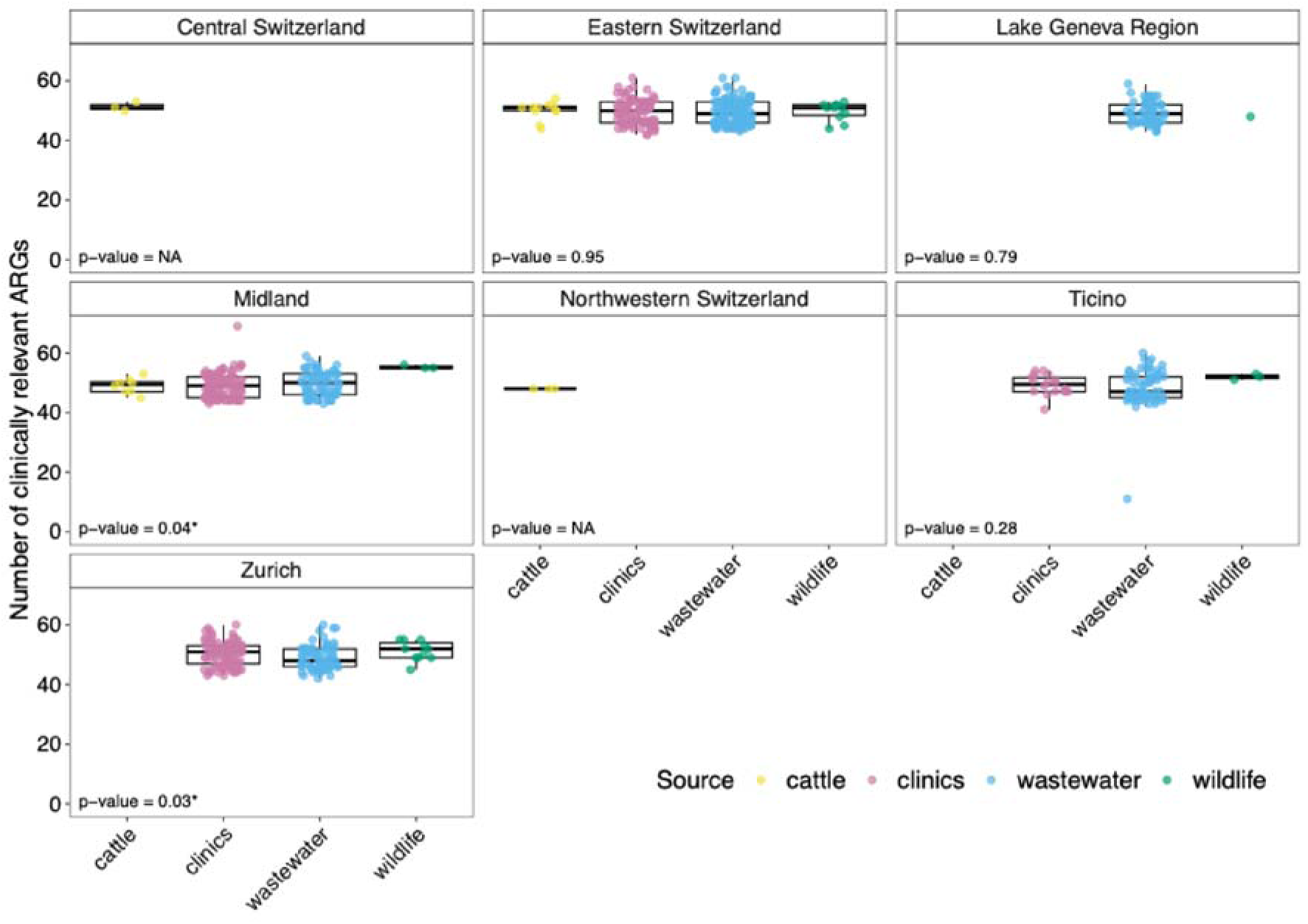
Regional variation in antibiotic resistance genes (ARGs) distribution across clinical, cattle, wastewater, and wildlife sources in Switzerland. Boxplots show the number of ARGs detected per isolate from clinics (pink), cattle (gold), wildlife (green), and wastewater (blue) across regions of Switzerland. Each panel represents a distinct region, and each boxplot reflects the distribution of ARG counts within a specific source in that region. Only sources with at least three isolates per region are included. *P*-values from the Kruskal-Wallis test are indicated in the bottom left corner of each panel. Significant differences (p-value < 0.05) are marked with *. When *p*-value = NA, the test was not performed due to the presence of only one source category in that region.

**Extended Data Fig. 3:**
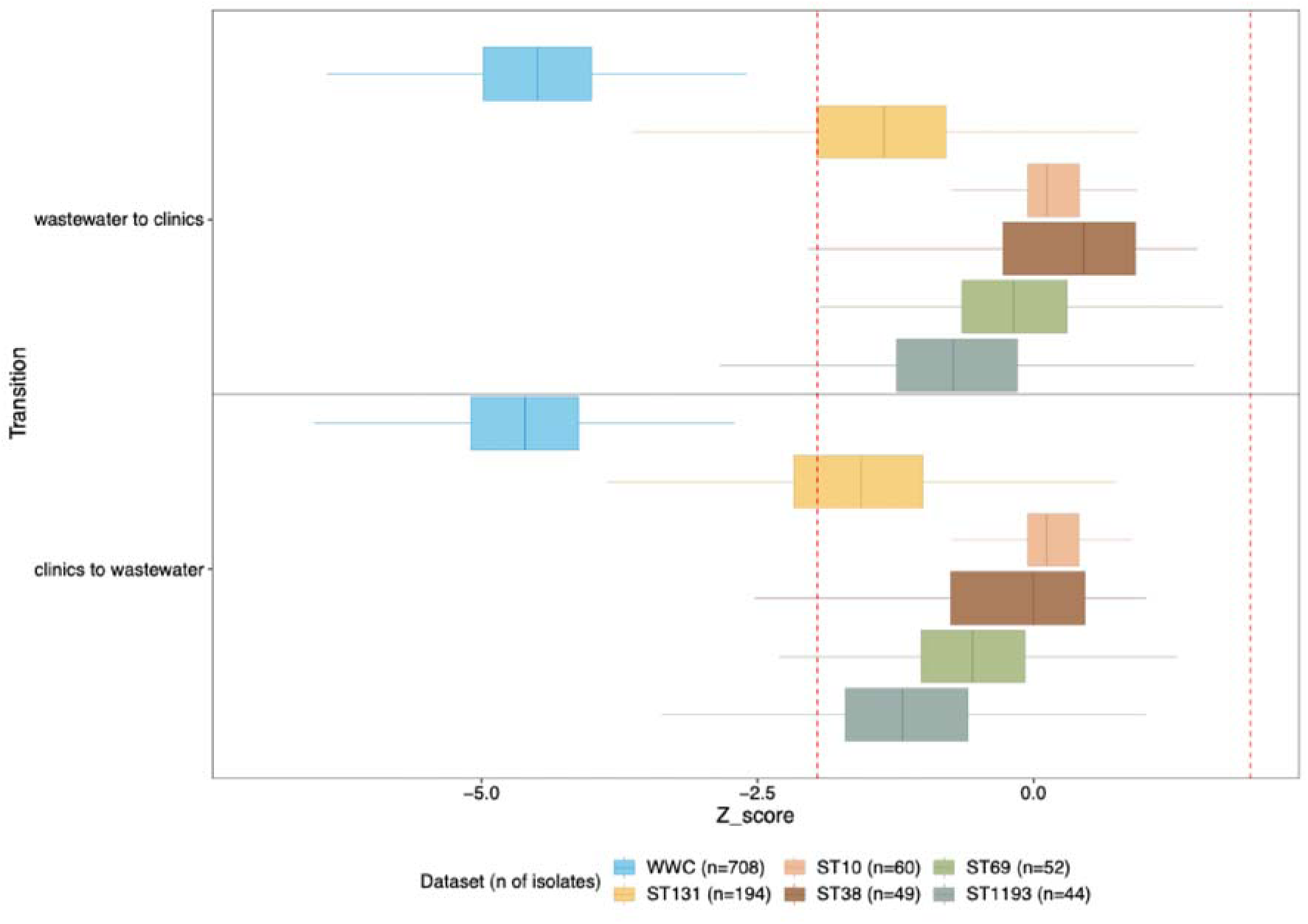
Z-score distributions of collection source transitions across 1,000 bootstrap phylogenies for wastewater and clinical isolates (WWC). Transitions between collection sources (clinics, wastewater) were inferred using maximum parsimony ancestral state reconstruction across 1,000 bootstrap phylogenetic trees, considering only 708 isolates from wastewater and clinical sources. For each tree, tip states were randomized 1,000 times (preserving source frequencies) to generate a null distribution of transition counts. Z-scores represent standardized deviations of observed transitions relative to the null. Each boxplot shows Z-score distributions per transition type, stratified by dataset subset (WWC-only full dataset and the five dominant sequence types: ST131, ST10, ST38, ST69, ST1193, all restricted to WWC isolates). The legend indicates the number of isolates retained in each subset. Red dashed lines mark ±1.96, corresponding to the central 95% confidence interval under the null model. Transitions were reconstructed using the *acctran* and *ancestral.pars* functions from the ape package (v5.7.1) in R (v4.1.2). Positive Z-scores indicate transitions occurring more frequently than expected by chance; negative Z-scores indicate less frequent transitions.

## Notes

### Competing Interest Statement

The authors have declared no competing interest.

### Author Declarations

In January, 2023, Repubblica e Cantone Ticino, Dipartimento della sanita e della socialite, Divisione della salute pubblica, Ufficio di sanita stated that Ethics Committee approval for the project is not needed because the bacterial strains are anonymized, so the research does not fall in the field of application of Human research Act Art. 2 and 3, and does not need approval.

