## Supplemental Material for "From clinics to sewers: leveraging environmental surveillance and whole genome sequencing to inform transmission of ESBL-*Escherichia coli* in Switzerland"

**SUPPLEMENTARY MATERIAL**

**Supplementary figures**

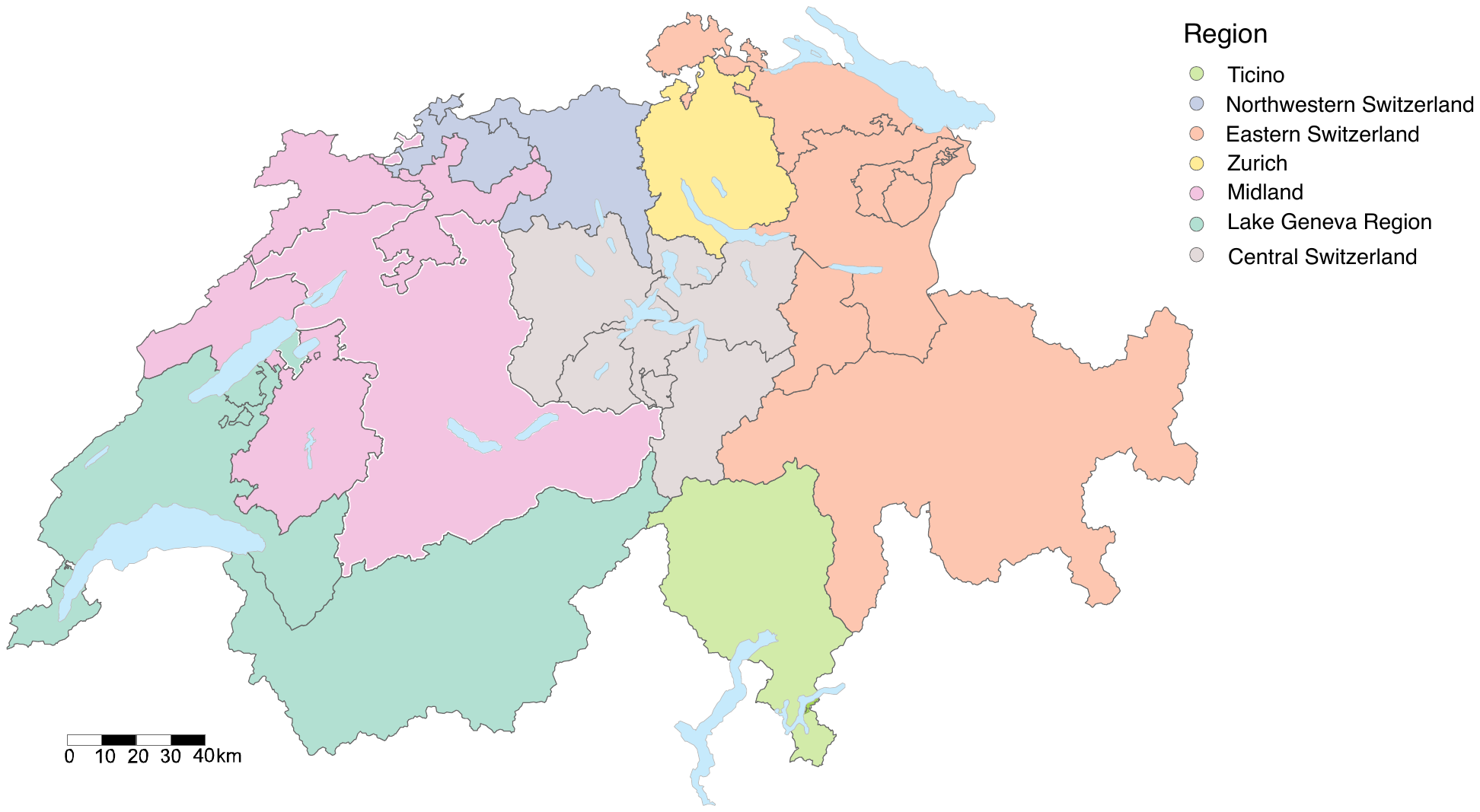
**Fig. S1:** **Regions of Switzerland.**

Map displaying the seven major regions of Switzerland as defined on the Swiss Federal Administration website (<https://www.admin.ch/gov/de/start/dokumentation/medienmitteilungen.msg-id-10585.html>). The regions are Central Switzerland, Eastern Switzerland, Lake Geneva Region, Midland, Northwestern Switzerland, Ticino, and Zurich. The map was generated in R (v4.1.1) and modified in Inkscape (v1.1.1).

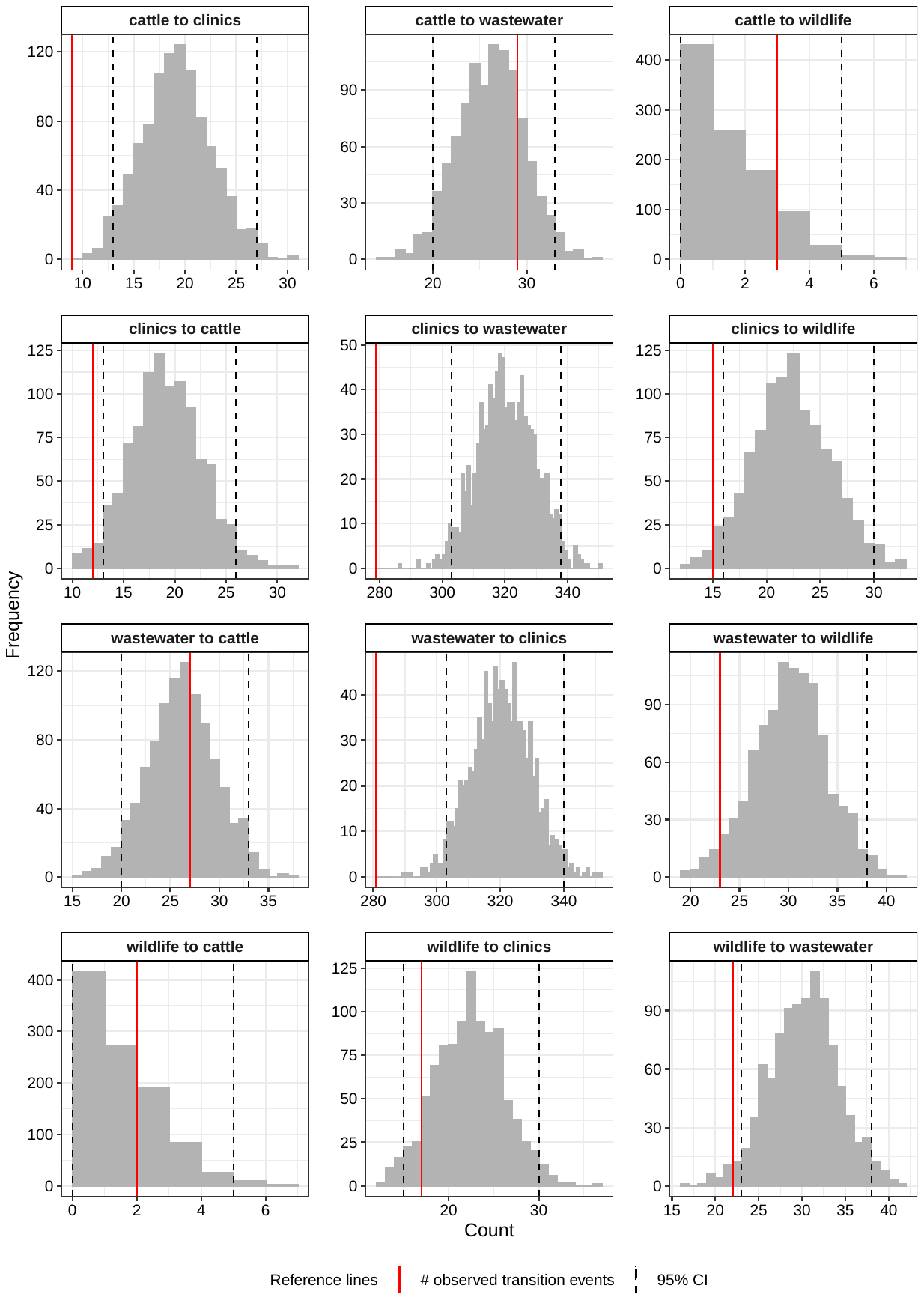

**Fig. S2: Distribution of observed and randomized transition counts on the consensus phylogeny.** Transitions between collection sources (clinics, wastewater, wildlife, cattle) were inferred using maximum parsimony ancestral state reconstruction on the consensus maximum likelihood phylogenetic tree of 762 isolates. For each transition type, tip states were randomized 1,000 times (preserving source frequencies) to generate null distributions of transition counts. Each facet shows the distribution of transition counts from the null model (histogram) for a specific transition type. Solid red lines indicate the observed number of transitions; dashed black lines mark the central 95% confidence interval from the null model. Transitions were reconstructed using the *acctran* and *ancestral.pars* functions from the ape package (v5.7.1) in R (v4.1.2).

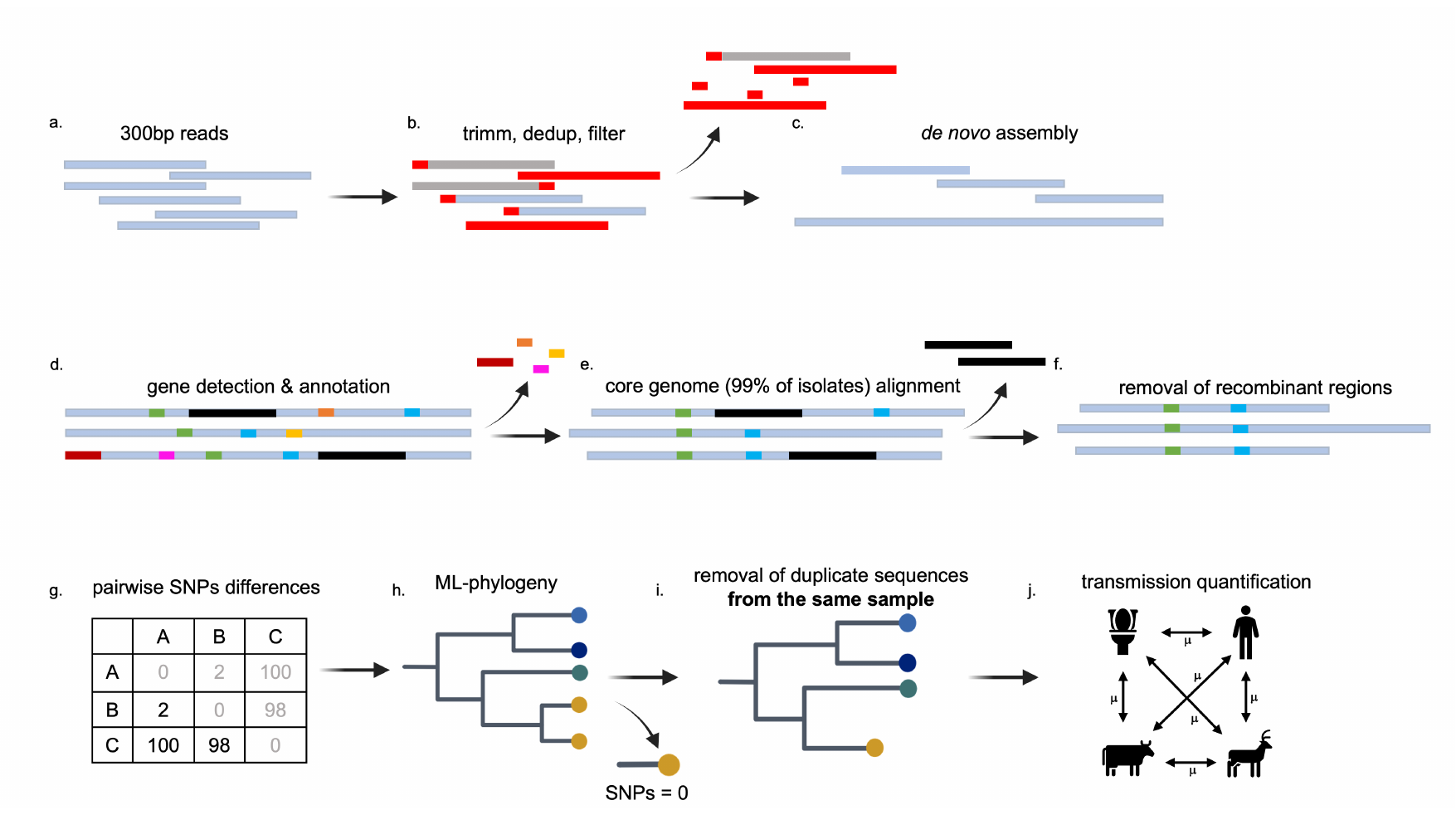
**Fig. S3: Bioinformatic analysis pipeline for ESBL-*E. coli*.**

This figure illustrates the bioinformatics workflow used for the analysis of ESBL-producing *Escherichia coli* isolates, implemented through a Snakemake workflow. The pipeline consists of multiple steps: (a) processing 300bp sequencing reads, (b) quality trimming, deduplication, and filtering, followed by (c) de novo genome assembly. (d) Gene detection and annotation were performed, while (e) core genome alignment (99% of isolates) was constructed. (f) Recombinant regions were removed to ensure accurate phylogenetic reconstruction. (g) Pairwise SNP differences were computed, and (h) a maximum-likelihood (ML) phylogenetic tree was generated. (i) Identical sequences from the same sample were removed to prevent redundancy, and (j) transmission events between compartments were quantified.

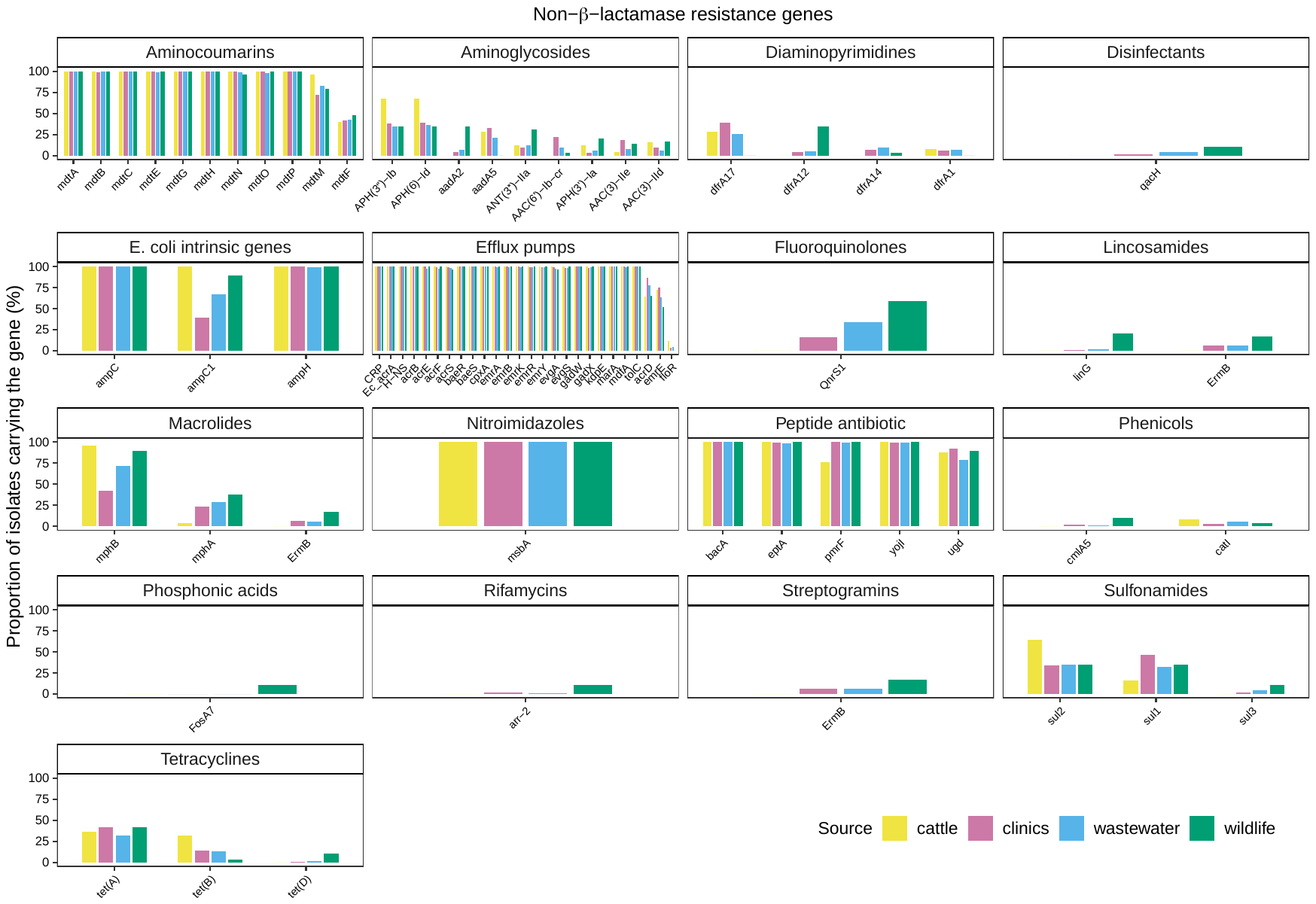
**Fig. S4: Distribution of antibiotic resistance genes across sources in ESBL-producing *E. coli* isolates.**

This figure shows the proportion of ESBL-*E. coli* isolates carrying genes associated with various antibiotic resistance classes, based on annotations from the Comprehensive Antibiotic Resistance Database (CARD). All resistance genes, excluding β-lactamases, that were detected in at least 5% of isolates from any source are displayed. Each panel represents a different antibiotic class, with colors indicating the sources: clinics (pink), cattle (gold), wildlife (green), and wastewater (blue). Antibiotic resistance classes include aminocoumarins, aminoglycosides, diaminopyrimidines, fluoroquinolones, lincosamides, macrolides, nitroimidazoles, peptides, phenicols, phosphonic acids, rifamycins, sulfonamides, tetracyclines, streptogramins, and disinfectants. When a gene is annotated as conferring resistance to multiple antibiotic classes, it is displayed in all relevant facets. Efflux pumps represent genes conferring resistance to multiple antibiotic classes by actively exporting antibiotics from bacterial cells. Each bar shows the proportion of isolates from a specific source carrying the corresponding gene.

**SUPPLEMENTARY TABLES**

**Table S1:** Comprehensive summary of ESBL-*E. coli* sequencing outcomes, quality filtering, control types by strain, and resequencing success across batches.

**Table S2:** Summary report of the quality assessment of the genome assemblies conducted using QUAST v5.0.2. The QUAST report, generated as an Excel file, includes various metrics for evaluating assembly quality, such as the number of contigs, total length, N50, L50, and GC content. The report provides a comprehensive comparison of assembly statistics to assess the completeness, accuracy, and overall quality of the genome assemblies.

**Table S3:** Metadata of extended-spectrum β-lactamase (ESBL)-producing *Escherichia coli* isolates collected from wastewater across various regions in Switzerland. The table includes sample identifiers, geographic location (city and region), collection date, detailed source, phylogenetic group, and multi-locus sequence type (ST). These isolates were analyzed to assess genetic diversity, phylogeny, and potential transmission dynamics of ESBL-producing *E. coli* within a One Health framework.

**Table S4:** Distribution of extended-spectrum β-lactamase (ESBL)-producing *Escherichia coli* isolates across nine phylogroups, determined using the EzClermont protocol, stratified by source.

**Table S5:** Distribution of extended-spectrum β-lactamase (ESBL)-producing *Escherichia coli* isolates by sequence type (ST), identified by the analysis of seven housekeeping genes of the multilocus sequence typing (MLST) Achtman scheme using MLST v.2.16.1 (<https://github.com/tseemann/mlst>). The table includes the number and percentage of isolates for each ST.

**Table S6:** Distribution of β-lactamase genes among extended-spectrum β-lactamase (ESBL)-producing *Escherichia coli* isolates from different sources. The table lists detected β-lactamase genes, their presence across sources (clinics, cattle, wastewater, and wildlife), the percentage of isolates carrying each gene, and the corresponding number of isolates. Gene detection was performed by blasting assembled genomes against the Comprehensive Antibiotic Resistance Database (CARD, accessed 4th November 2023) using Abricate v1.0.1.

**Table S7:** Statistical analysis of differences in antibiotic resistance gene (ARG) number among sources within different regions of Switzerland. The first part of the table presents the results of the Kruskal-Wallis test, indicating the chi-square (*χ^2^*) and p-value for each region: Eastern Switzerland, Midland, Lake Geneva Region, Ticino, and Zurich. The second part of the table shows the results of Dunn’s test with Bonferroni adjustment for pairwise comparisons across all tests, including the Z-score, p-value, and adjusted p-value for each comparison between sources (clinics, cattle, wastewater, and wildlife). Significant comparisons are highlighted in green to indicate regions where sources differ significantly in ARGs number.

**Table S8:** Isolate pairs of extended-spectrum β-lactamase (ESBL)-producing Escherichia coli that were genetically identical (0 SNPs). For each pair, the source compartment, sampling date, and region are shown. The table includes both within-compartment and between-compartment matches; rows shaded in green represent isolate pairs originating from different compartments.

**Table S9:** Significance summary of transition frequencies across 1,000 bootstrap phylogenies for the full dataset and the five most dominant sequence types (ST131, ST10, ST38, ST69, ST1193). The table reports the number of bootstrap trees in which the observed number of transitions between sources fell below, within, or above the central 95% confidence interval of the null distribution generated by 1,000 random permutations. Transitions with counts below the 95% CI (columns "Below 95% CI") or above the 95% CI (columns "Above 95% CI") indicate significant deviation from random expectations at the 0.05 level.

**Table S10:** Significance summary of transition frequencies across 1,000 bootstrap phylogenies based on isolates from wastewater and clinical sources only (WWC). The table reports the number of bootstrap trees in which observed transitions between wastewater and clinics fell below, within, or above the central 95% confidence interval (CI) of the null distribution from 1,000 random permutations. Results are shown for the full WWC phylogeny and for the five most dominant sequence types (ST131, ST10, ST38, ST69, ST1193). Transitions falling below the 95% CI (column "Below 95% CI") indicate significantly fewer transitions than expected under the null model.

**Table S11:** Identical ESBL-*E. coli* isolates (0 SNPs) originating from the same sample and collapsed into a single representative. For each case, the retained isolate is listed alongside the collapsed identical sequences, source compartment, and collection date. This includes, for example, multiple clonal isolates obtained from the same wildlife or livestock fecal sample, or from the same wastewater sample.
